# Statin use is associated with reduction in risk of *de novo* primary sclerosing cholangitis among patients with inflammatory bowel disease: A National Database Study

**DOI:** 10.1101/2024.09.17.24313852

**Authors:** Chiraag Kulkarni, John Gubatan, C. William Pike, Gavin Hui, Saurabh Gombar, Jennifer Kramer, Michael L. Jackson, Aparna Goel, John Vierling, George Cholankeril, Sidhartha R. Sinha

**Author notes:** **Correspondence:** Sidhartha R. Sinha, MD, Division of Gastroenterology and Hepatology, Stanford University School of Medicine.

## Abstract

**Background and Aims:** Primary sclerosing cholangitis (PSC) is a liver disease without medical therapy that significantly increases the risk of malignancies, acute cholangitis and cirrhosis. Approximately 2-7% of patients with inflammatory bowel disease (IBD) develop PSC. No drug has been shown to prevent *de novo* PSC in patients with IBD. Statin use, however, has been associated with increased liver transplant-free survival. Yet, the impact of statins on development of new PSC among those with IBD is unknown.

**Methods:** We conducted a retrospective cohort study of patients diagnosed with IBD with and without statin exposure using a large, representative national database. Patients were followed from the date of diagnosis of IBD. Patient demographics, co-morbidities, medications, and type of IBD were extracted. Unmatched and propensity score-matched Cox regression analyses were performed.

**Results:** Our analysis included 33,813 patients with IBD of whom 8,813 were exposed to statins. PSC developed in 181 patients over a median follow-up of 3.7 years. Only nine patients (0.1%) who were exposed to statins developed PSC compared to 173 (0.69%) in the non-exposed population. In propensity score-matched analysis, statin therapy was associated with an 86% lower risk of developing PSC (HR 0.14; 95% CI 0.06-0.33, p<0.001). The findings were consistent when accounting for unmeasured confounders (E-value) in sensitivity analyses, including stratification by age group, duration of statin use, and type of statin.

**Conclusion:** In a propensity score-matched analysis, statin use was associated with an 86% risk reduction in new PSC diagnosis among patients with known IBD, suggesting a potential benefit as a prophylactic agent. These findings warrant prospective validation.

## Introduction

Primary sclerosing cholangitis (PSC) is a progressive, cholestatic liver disease that has no approved disease-modifying medicine for prevention or treatment. Patients with PSC have significantly elevated risks of multiple malignancies (e.g., cholangiocarcinoma, colorectal carcinoma, hepatocellular carcinoma), recurrent acute cholangitis, and cirrhosis with decompensation requiring liver transplantation. PSC is characterized by injury to cholangiocytes, resulting in foci of biliary inflammation and stricturing fibrosis^1,2^. The median age for diagnosis of PSC among patients with inflammatory bowel disease (IBD) is approximately 42 years^3,4^. While approximately 80% of patients with PSC have IBD, only 2-7% of patients with IBD have PSC^2^.

There is growing evidence that statins, which can alter serum bile acid profiles and modulate chronic inflammation, are beneficial in chronic liver diseases and IBD^5-11^. PSC is characterized by dysbiosis and alterations in bile acid (BA) profile^5^. An increased ratio of primary bile acids (PBAs) to secondary bile acid (SBAs) was predictive for hepatic decompensation, suggesting that alteration of BA profiles is a potential therapeutic target^6^. This same BA imbalance also contributes to pouchitis, a complication of IBD surgery that occurs more commonly in patients with PSC^7^. Statins are known to increase SBA levels and, thus, might correct BA imbalance in PSC^8^.

In preclinical models of chronic liver disease, statins reduce paracrine activation of hepatic stellate cells and attenuate oxidative stress, thereby reducing hepatic fibrosis^9^. In retrospective clinical studies, patients with chronic liver disease who used statins had a decreased rate of progression to cirrhosis^10,11^. Furthermore, statin use in patients with compensated cirrhosis decreased the rate of progression to decompensated cirrhosis and death^10,11^. Additionally, statins have been associated with diminish chronic inflammation in IBD, reducing the need for colectomy by approximately 50% among patients with ulcerative colitis (UC)^12^. Finally, retrospective studies in PSC patients showed an association between statin use and longer transplant-free survival and decreased all-cause mortality^13^.

However, the impact of statin use on the development of PSC in patients with IBD remains unknown. Thus, the objective of the study was to determine the impact of statin use on the risk of developing *de novo* PSC among patients with preexisting IBD.

## Materials and Methods

### Data Source

We used a representative national database, EVERSANA, consisting of over 120 million de-identified patients in the US. The data are generated at the point of care and entered into multiple electronic medical record (EMR) systems. The data is sourced from >2,000 outpatient/ambulatory health centers, >500 hospitals, >30 health systems (including academic medical centers) and >50 unique EMR platform providers (e.g. EPIC, CERNER, etc.).

### Study Population and Outcome

We used a new-user design among patients with newly diagnosed IBD comparing new statin users with new users of any non-statin medication. IBD was defined by the first mention in the database by an International Classification of Disease 10 (ICD-10) code corresponding to ulcerative colitis (UC) or Crohn’s disease (CD). For patients with CD, we were unable to determine the specific location of disease. We eliminated anyone with a prior ICD 9 code for UC or CD and eliminated anyone with more than 1 measurement of fecal calprotectin greater than or equal to 250 mcg/g to ensure that the first ICD-10 code for either UC or CD truly represented a new diagnosis of IBD, rather than first recording of a diagnostic code. We validated this definition of new diagnosis of IBD using Stanford Research Repository (STARR) to ensure high classification performance, judged against the gold standard of manual physician review. Physician determination was made by the evaluation of two separate physicians, and if there was disagreement the tie was broken by a third physician.

Patients were followed from the first mention of IBD in the database, starting on Nov 1, 2018, until Oct 31, 2023. Patients could not have a diagnosis of PSC prior to cohort entry. The outcome was a new diagnosis of PSC by ICD-10 code (K83.01) without prior ICD-9 coding for cholangitis, or 3 or more previous values of alkaline phosphatase greater than 1.5 times the upper limit of normal. This was done to minimize inclusion of patients with PSC and ensure the first ICD-10 code for PSC was truly a new diagnosis. This definition of new PSC was also validated in STARR, judging against manual review by physicians. Full performance characteristics for both definitions are included in the supplementary materials

Further, all patients were required to have sufficient history as determined by presence of an EHR entry within the last 90 days. They were also required to have at least six months of follow-up from the time of IBD diagnosis to allow for a latency-time window and minimize detection bias.

### Exposure definition

Statin use was identified by Anatomical Therapeutic Chemical (ATC) Classification System (code C10AA). The comparison group was the start of any non-statin drug. Within 90 days of IBD diagnosis, patients were required to have exposure to statin or other control medication. We took care to avoid misclassification of immortal time in the statin exposed group by categorizing time between IBD diagnosis and statin prescription as untreated. A new diagnosis of PSC was identified by ICD-10 code (K83.01). Patients were considered at risk of PSC development beginning on day 1 (time of IBD dx) in the primary analysis. Given uncertainly regarding the optimal latency time for the effect of statins, sensitivity analyses with lag times were performed.

### Statistical analysis

Descriptive statistics were applied to the demographic and clinical characteristics of patients with and without statin use. Student’s t-test was used to compare continuous variables and the chi-squared test to compare categorical variables.

We considered a wide range of potential cofounders, including age, sex, race/ethnicity and multiple comorbidities. Accordingly, we performed unmatched, basic matched (age, sex, race/ethnicity), and high dimensional propensity score (PS) matched analyses. Matching was done 1:1, with caliper distance of 0.2. Distance tolerance to be considered for exact matching was 0.01. PS matching was performed using all components of the Charlson Comorbidity Index (solid tumor, leukemia, lymphoma, diabetes, congestive heart failure (CHF), myocardial infarction (MI), peripheral vascular disease (PVD), chronic obstructive pulmonary disease (COPD), cerebrovascular disease (CVD), dementia, hemiplegia, liver disease, chronic kidney disease (CKD), peptic ulcer disease (PUD), rheumatic disease, and HIV), year of entry into the cohort, medications, age, sex and race/ethnicity. Standardized mean difference was calculated for each variable. Propensity score (PS) distributions were checked to ensure adequate overlap.

In primary analysis, we calculated unmatched, basic matched, and PS adjusted hazard ratio of new onset PSC using Cox regression among patients with IBD, comparing statin users versus non-users. As noted above, we took precautions to systematically limit the possibility of immortal time bias by mandating start time of statin and a comparator medication to be within 90 days of IBD diagnosis, and further ensure there was no misclassification of immortal time among statin-exposed patient. To account for possible unmeasured confounding, E-values were calculated. E-value measures, on a ratio scale, the strength of unmeasured confounding that would have to be associated with both the treatment and outcome to negate the observed association. For example, a E-value of 5.0 indicates than an unmeasured confounder would have to be associated with both the treatment (statins) and the outcome (new onset PSC) by a risk ratio of 5.0 beyond all measured confounders to explain away the observed association, and weaker confounding would not do so. In this study, E-values are reported for the high-dimensional propensity score matched hazard ratios (HR), where residual confounding is likely to be relatively low^14,15^. We performed pre-specified secondary and sensitivity analyses to assess the robustness of our findings. First, we evaluated the possibility of type specific effect of statins, comparing the impact of hydrophilic (rosuvastatin and pravastatin) and lipophilic (simvastatin, fluvastatin, lovastatin, and pitavastatin) statins separately. Second, we assessed the impact of statin therapy on patients less than 50 years old, which is the most common age group for newly diagnosed PSC patients. Third, given the male predominance among patients with PSC, we determined the impact of sex. Fourth, given the uncertainty regarding the optimal latency time window, we redid the primary analysis with a lag time of 12 months. Fifth, we calculated the impact of duration of statin therapy, comparing patients with less than 2 years of exposure to those with greater than 2 years of therapy. Finally, we evaluated if there was a difference in *de novo* PSC by type of IBD, namely ulcerative colitis (UC) or Crohn’s disease (CD).

### Ethical Considerations

The study was deemed exempt from review by the Institutional Review Board of Stanford University because of its use of retrospective, deidentified data.

## Results

### Baseline Clinical Characteristics

The study population included 33,813 patients with newly diagnosed IBD of which 8,813 were prescribed statins from 2018-2023. Over the follow-up period, there were 173 new cases of PSC among patients not prescribed statins (0.69%) and 9 cases among patients prescribed statins (0.1%). The cohort was 51.6% Caucasian and mean age for the cohort was 51.1 years. The population was 57.2% female. In the unmatched cohort, patients prescribed statins were, on average, older (62.9 years versus 46.9 years); they also had higher rates of diabetes, CHF, MI, PVD, COPD, cerebrovascular disease and renal disease. Clinical characteristics comparing patients with IBD who did and did not take statins before and after PS matching are shown in **Table 1**. The PS matched cohort had 5,627 patients in each arm, and there were no major differences in patient composition.

**Table 1.** Demographics and Clinical Characteristics Before and After Propensity Score Matching.

|  | No statin use<br>(unmatched) | Statin use<br>(unmatched) | No statin use (PS<br>matched) | Statin use<br>(PS<br>matched) |
| --- | --- | --- | --- | --- |
| N | 25,000 | 8,813 | 5,627 | 5,627 |
| Female (%) | 14,673<br>(58.7%) | 4,687<br>(53.2%) | 3,086<br>(54.8%) | 3,096<br>(55.0%) |
| Mean age (sd) | 46.9 (15.9) | 62.9 (9.6) | 62.1 (10.6) | 61.2 (9.7) |
| Race (%) |  |  |  |  |
| White | 13,501<br>(54.0%) | 3,950<br>(44.8%) | 2,853<br>(50.7%) | 2,810<br>(49.9%) |
| Black | 1,330 (5.3%) | 333 (3.8%) | 230 (4.1%) | 241 (4.3%) |
| Asian | 405 (1.6%) | 71 (0.8%) | 61 (1.1%) | 46 (0.8%) |
| Other | 9,764 (39.1%) | 4,459<br>(50.6%) | 2,482<br>(44.1%) | 2,530<br>(45.0%) |
| Hispanic (%) | 942 (3.8%) | 525 (6%) | 212 (3.8%) | 287 (5.1%) |
| Comorbidity score (sd) | 1.6 (2.2) | 4 (3.2) | 3.5 (2.9) | 3.4 (2.7) |
| Malignancy | 1,507 (6.0%) | 978 (11.1%) | 780 (13.9%) | 486 (8.6%) |
| Metastatic Solid Tumor | 395 (1.6%) | 211 (2.4%) | 232 (4.1%) | 84 (1.5%) |
| Diabetes | 1,710 (6.8%) | 3,140<br>(35.6%) | 852 (15.1%) | 1,649<br>(29.3%) |
| Complicated Diabetes | 480 (1.9%) | 1,269<br>(14.4%) | 310 (5.5%) | 551 (9.8%) |
| CHF | 528 (2.1%) | 999 (11.3%) | 300 (5.3%) | 475 (8.4%) |
| MI | 276 (1.1%) | 666 (7.6%) | 138 (2.5%) | 364 (6.5%) |
| PVD | 745 (3.0%) | 1183<br>(13.4%) | 397 (7.1%) | 581 (10.3%) |
| COPD | 4,028 (16.1%) | 2,348<br>(26.6%) | 1,348<br>(24.0%) | 1,345<br>(23.9%) |
| CVD | 642 (2.6%) | 1,153<br>(13.1%) | 340 (6.0%) | 564 (10.0%) |
| Dementia | 72 (0.3%) | 138 (1.6%) | 45 (0.8%) | 55 (0.98%) |
| Hemiplegia | 115 (0.5%) | 148 (1.7%) | 47 (0.8%) | 71 (1.3%) |
| Mild Liver Disease | 1,389 (5.6%) | 783 (8.9%) | 567 (10.1%) | 421 (7.5%) |
| Severe Liver Disease | 184 (0.7%) | 99 (1.1%) | 110 (2.0%) | 48 (0.9%) |
| Renal Disease | 849 (3.4%) | 1,180<br>(13.4%) | 528 (9.38%) | 531 (9.4%) |
| PUD | 567 (2.3%) | 308 (3.5%) | 197 (3.5%) | 160 (2.8%) |
| Rheumatic Disease | 851 (3.4%) | 446 (5.1%) | 325 (5.8%) | 256 (4.6%) |
| HIV | 123 (0.5%) | 51 (0.6%) | 54 (1.0%) | 17 (0.3%) |
| Outcome |  |  |  |  |
| PSC | 173 (0.69%) | 9 (0.1%) | 41 (0.73%) | 6 (0.11%) |

### Statin therapy is associated with lower risk of *de novo* PSC among patients with IBD

Statin therapy was associated with a significant reduction in the diagnosis of new-onset PSC among patients with IBD in unmatched (HR 0.16, 95% CI 0.08-0.31, E-value 6.0), basic matched (HR 0.16, 95% CI 0.08-0.33, E-value 5.5), and high-dimensional PS matched analysis (HR 0.14, 95% CI 0.06-0.33, E-value 5.5) using a Cox model. In the PS matched group, the incidence of new-onset PSC among statin users was 0.11% compared to 0.73% among patients on a comparator medication (**Figure 1**).

**Figure 1.**
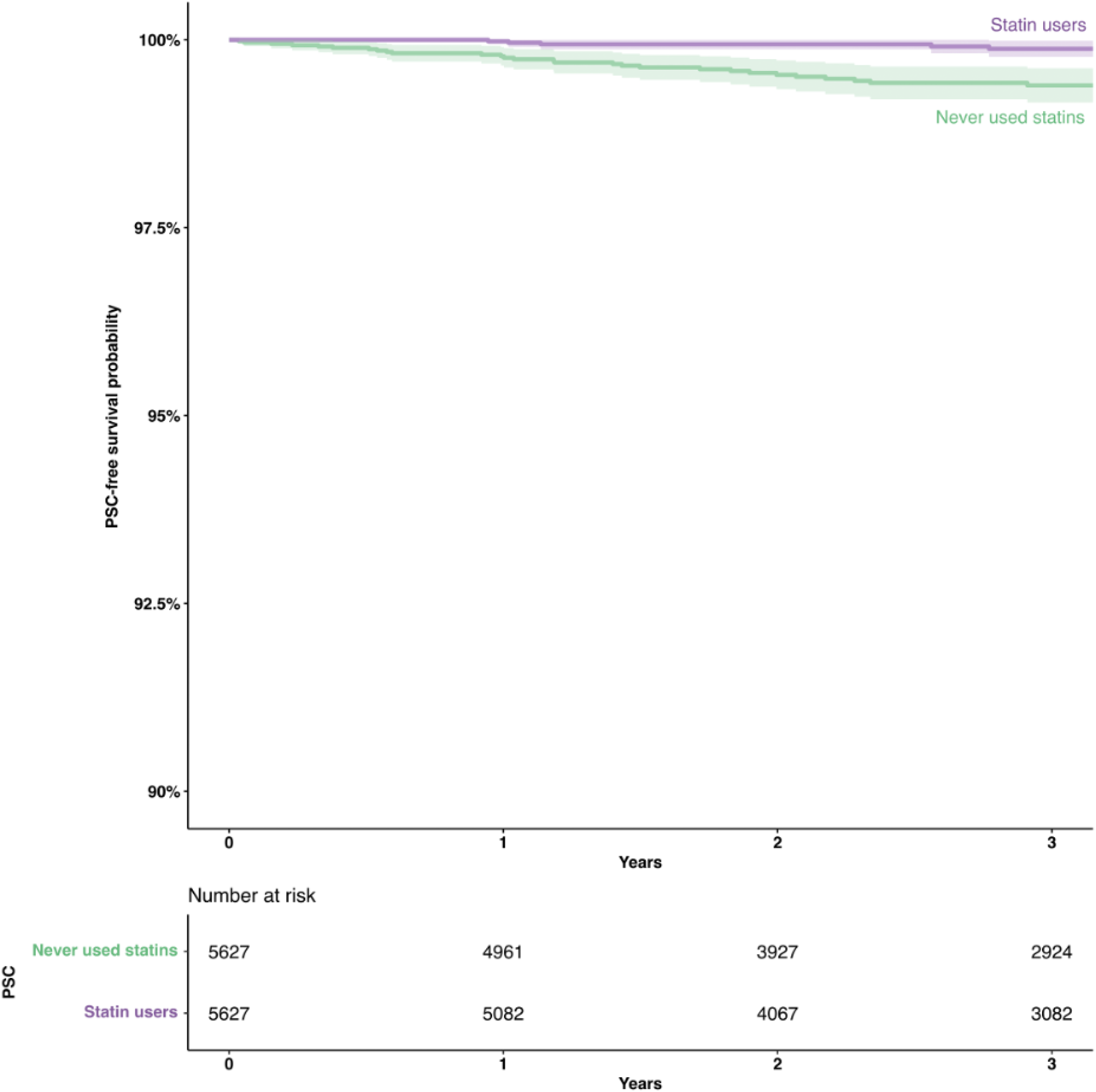
Propensity score matched hazard of PSC among patients with IBD, among statin users and non-users who were prescribed a comparator medication

The protective association against *de novo* PSC among patients with IBD was independent of the class of statin. Namely, the magnitude of the impact observed in PS matched analysis was similar for both lipophilic statins (HR 0.08, 95% CI 0.02-0.33, E-value 5.6) and hydrophilic statins (HR 0.07, 95% CI 0.01-0.50, E-value 3.4).

The average age of onset of PSC among patients with IBD is approximately 40-42 years^3,4^. Therefore, in addition to the primary PS matched analysis we also evaluated the impact of statin therapy in the sub-group of patients less that 50 years of age. In the PS matched analysis of 1,776 patients, statin users had a 90% risk reduction in new-onset PSC (HR 0.1, 95% CI 0.01-0.83, E-value 2.1).

We next evaluated whether sex influenced the impact of statin therapy on PSC development. Among male patients with IBD on statin therapy (n=4,126), there was a lower risk of new PSC compared to non-users in PS matched analysis (HR 0.22, 95% CI 0.08-0.59, E-value 2.8). Similarly, among female patients with IBD on statin therapy (n=4,687), there was a risk reduction of similar magnitude (HR 0.16, 95% CI 0.04-0.70, E-value 2.2).

Patients included in the primary analysis were considered at risk for PSC starting on day 1 of follow-up. We repeated the analysis using a lag time of 12 months, given uncertainty about the optimal latency period. Using a lag time of 12 months, we observed a 79% risk reduction in the odds of new PSC among patients with IBD in PS matched analysis (HR 0.21, 95% CI 0.09-0.52, E-value 3.3). We also evaluated the impact of duration of therapy on the risk of *de novo* PSC, comparing patients with short-term statin exposure (0-2 years) to patients with long-term exposure (> 2 years). Among patients with short-term exposure (0-2 years, n=3,233), there was a 76% risk reduction in new PSC (HR 0.24, 95% CI 0.09-0.64, E-value 2.5); among patients with long-term exposure (> 2 years, n=5,996) there was a 75% risk reduction in new PSC (HR 0.25, 95% CI 0.07-0.42, E-value 3.2).

We determined the risk of *de novo* PSC among patients with IBD evaluating hyperlipidemia as a risk factor in patients not on statin therapy. When comparing patients with IBD and hyperlipidemia without statin exposure to patients with IBD without hyperlipidemia without statin exposure, there was no difference in *de novo* PSC (n=35,242, HR 1.01, 95% CI 0.73-1.40).

We evaluated whether the type of IBD impacted the risk of new-onset PSC. Among patients with UC on statin therapy (n=5,547), there was a 79% reduction in new-onset PSC compared to non-users (HR 0.21, 95% CI 0.08-0.56, E-value 3.0). Similarly, among patients with CD taking a statin (n=2,817) there was an 85% reduction in new PSC compared to non-users (HR 0.15, 95% CI 0.03-0.65, E-value 2.4). PS matched results from the primary, secondary and sensitivity analyses are summarized in a forest plot (**Figure 2**).

**Figure 2.**
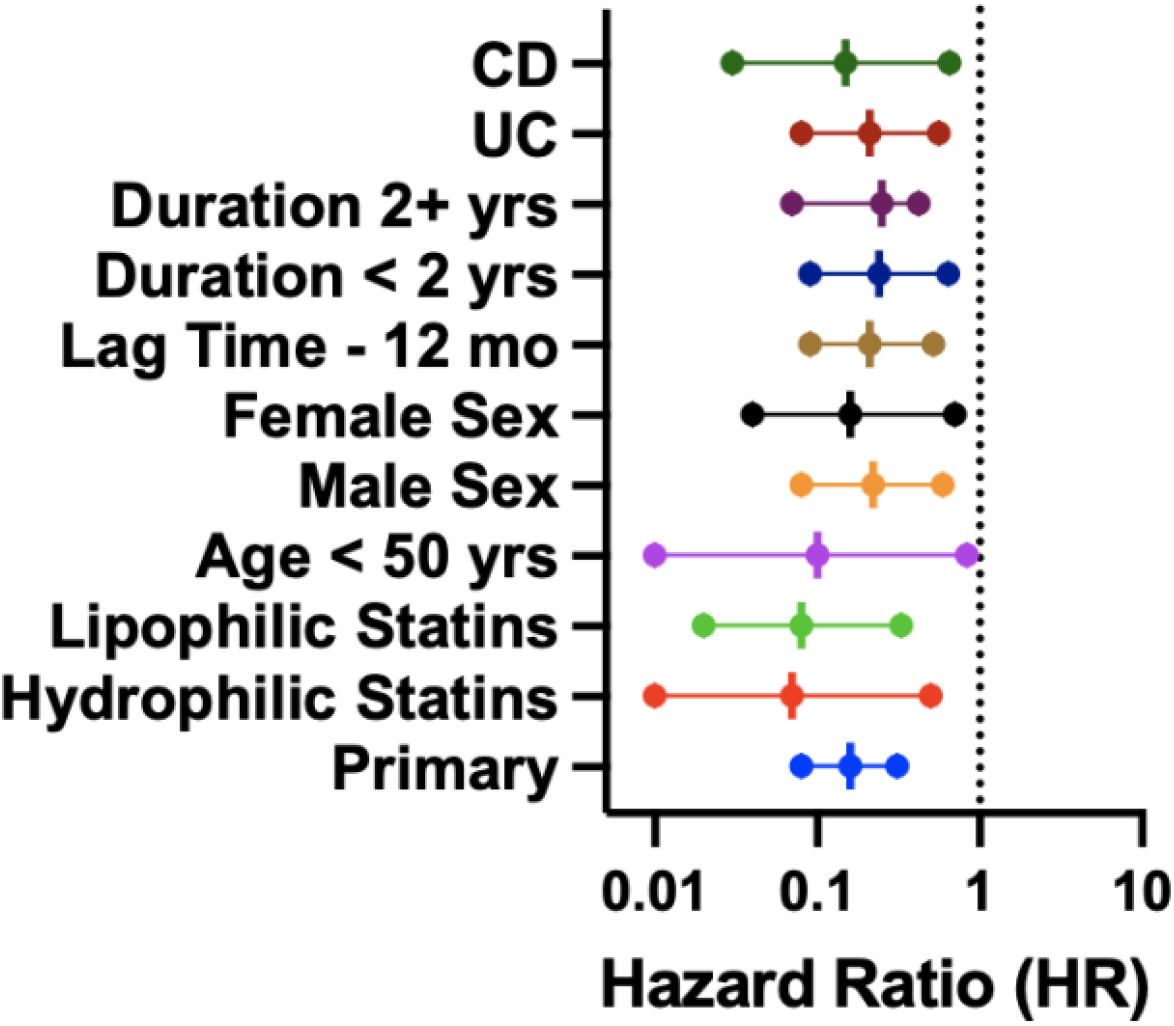
Forest plot summarizing primary, secondary and sensitivity analyses

## Discussion

PSC is a progressive, morbid, cholestatic liver disease that is strongly associated with IBD. To date, there is no effective medical therapy for the treatment or prevention of PSC^16,17^. This is the first study showing that statin therapy is associated with a profound reduction (86%) in the risk of developing new-onset PSC among patients with IBD in PS matched analysis of a large, representative national database. An E-value of 5.5 in PS matched suggests that the observed associated is highly unlikely to be due to residual confounding. Our results are consistent in the secondary and sensitivity analyses, which addressed the most likely potential sources of biases, strengthening our findings.

Statins are a safe, well-tolerated, and widely prescribed class of medication. In this study, we show the potential for statins to be used as a prophylactic agent for the prevention of PSC among patients with IBD. Patients with PSC are known to have dysbiosis and altered BA profile, and it is likely that statins affect the risk of new-onset PSC through modulation of inflammation, changes in the microbiome, and alterations in bile acid profile^8,12,18–20^. Patients with PSC have elevated PBAs compared to healthy controls and gut dysbiosis. Higher PBA levels have been shown to promote hepatic decompensation and intestinal inflammation^6^. Statins modulate BA profiles directly (via changes in cholesterol synthesis) and indirectly (potentially via modulation of the microbiome)^8,21,22^. Statin decrease PBAs while increasing SBAs, and the effects on BAs and the microbiome take place within weeks. We believe that statins may impact biliary inflammation similarly to intestinal inflammation in UC, where they alter UC transcriptomic signatures and are associated with a reduction in the risk of colectomy by 53%^12^. The results of our primary analysis and analysis of duration of use show the putative protective effect begins early, consistent with this posited mechanism. Taken together, these data provide a plausible biologic basis for statins preventing the onset of new PSC.

Our results are also in concordance with a growing body of literature showing the benefit of statins in chronic liver diseases, including PSC^9-11,23,24^. In both laboratory and clinical data, statins have been shown to slow progression of liver disease across a wide range of pathologies including steatotic liver disease, viral hepatitis, and cholestatic disorders^9–11,23,24^. Statins are associated with a reduction in the risk of mortality and cirrhosis among patients with PSC^13^. Our work extends these findings, suggesting that the potential for statins to be used in primary prevention of PSC among patients with IBD.

Our findings will require validation in randomized, controlled clinical studies. However, we believe that our current data do provide a basis for providers caring for patients with IBD to more aggressively address statin hesitancy among patients with IBD who already have an evidence-based indication for statin therapy. Recent data shows that just 33.6% of patients eligible for statin therapy by American College of Cardiology (ACC)/American Heart Association (AHA) cholesterol guidelines take statins^25^. Addressing statin hesitancy among patients with IBD will not only offer proven benefit in cardiovascular disease but may also provide protection against PSC. This is especially important given emerging data suggesting that patients with IBD are increased risk of cardiovascular disease^26^.

Even though our data show a significant 86% risk reduction in the onset of new PSC, the low incidence of PSC even among patients with IBD is a barrier to broad application and prospective validation. Assuming a 10-year risk of PSC among patients with new IBD to be between 2-3%, the number needed to treat (NNT) is between 50-60 patients. This is 2-to 3-fold higher than current standard of care for statin therapy in atherosclerotic cardiovascular disease (ASCVD)^27^. Identification of a higher risk population among patients with IBD is necessary, not only to bring in the NNT in line with standard of care for ASCVD, but also to make the sample size for a prospective trial feasible for a rare disease.

This study has several strengths. First, this study was performed using large representative national database and included nearly 34,000 patients. Second, we used high dimensional propensity score matching to control for a wide array of potential confounding variables; PS matched analysis and sensitivity analysis of the <50 years cohort support that the results showing the protective association for statin users are not due to survival bias. Third, we restricted the cohort to new users, eliminating biases associated with the inclusion of existing users. Fourth, all patients had adequate follow-up to minimize detection bias. Finally, our results remained highly consistent, both in direction and magnitude of association, across all sensitivity analyses. This study also has limitations. First, the database has limited capture of certain categories of data. It had limited reporting on dosage data, precluding analysis to determine dose effect. It also had limited reporting of IBD location (for example, small bowel CD vs. colonic CD vs ileocolonic CD), so we were unable to analyze if location of disease modulated the impact of statin therapy. Second, there is a risk of misclassification of exposure and outcome in patients who may have been prescribed a statin from a health system not reporting to the dataset as non-statin users; similarly, the first mention of IBD and/or PSC in the database may not have been the true date of diagnosis if the patient came from a health system not reporting to the dataset. However, given the database is aggregated from >2,000 healthcare systems, this risk is low; further, given our data showed a protective association with statin therapy, this would in fact bias against detection of a difference between the two groups. Third, patients who are prescribed statins have fundamental differences compared to non-users including older age and increased co-morbidities; however, to account for this we performed PS matching accounting for these differences. Fourth, due to the nature of a database study we are unable to assess adherence to statin therapy. Fifth, though we used a new-user design with a comparison medication, that medication did not necessarily have the same indication. To reduce the likelihood of confounding by indication, we performed PS matching to balance common indications between the two groups, namely myocardial infarction and cerebrovascular accident. Additionally, to address the possibility hyperlipidemia may impact the risk PSC, we compared the risk of *de novo* PSC in patients with IBD with hyperlipidemia not on statin therapy to patients with IBD without hyperlipidemia not on statin therapy and found no difference. Sixth, given the observational nature of this study, residual confounding remains possible though performing high-dimensional propensity score matching makes this much less likely. We calculated E-values to quantify the likelihood that residual confounding could explain our results; large E-values across multiple analyses suggest that it is unlikely that residual confounding could explain our results.

In conclusion, we identified, for the first time, that statins may prevent the onset of PSC among patients with IBD. Specifically, statins, which are a common, safe and well-tolerated class of medications, were associated with an 86% reduction in *de novo* onset of PSC. Given that PSC has significant morbidity and mortality and lacks effective medical therapy, the protective effects of statins warrant further study.

## Data Availability

All data produced in the present study are available upon reasonable request to the authors

